# Prevalence and Risk Factors of Diarrhea Among Children Under Five Years in Northern Kenya’s Drylands: A Longitudinal Study

**DOI:** 10.1101/2024.11.13.24317266

**Authors:** Bonventure Mwangi, Valerie L. Flax, Faith Thuita, Joshua D. Miller, Chessa Lutter, Dickson Amugsi, Estelle Sidze, Linda Adair, Esther Anono, Hazel Odhiambo, Stephen Ekiru, Gillian Chepkwony, Monica Ng’ang’a, Albert Webale, Elizabeth Kimani-Murage, Calistus Wilunda

## Abstract

Diarrhea is the third leading cause of malnutrition and mortality in children under five globally. However, a few studies have examined predictors of child diarrheal disease in arid or semi-arid regions of sub-Saharan Africa. This study aimed to assess the prevalence and risk factors of diarrhea among children under five in the drylands of Northern Kenya.

Data are from a longitudinal, population-based study of children younger than 36 months at recruitment (N = 1,211) and their caregivers in Turkana County, Kenya. Households were followed from May 2021 to September 2023, with survey data collected every four months across six waves. Caregivers reported on household conditions and observed episodes of diarrhea among index children in the two weeks before the survey. Trends in the prevalence of diarrhea were stratified by administrative zone, livelihood zone, and child sex. Risk factors of child diarrhea across survey waves were identified using multivariable generalized estimating equations.

Diarrhea prevalence decreased from 32.1% [95% confidence interval (CI): 28.3%-36.1%] at Wave 1 to 8.7% (95% CI: 6.3%-11.7%) at Wave 6. Risk factors for diarrhea included caregivers alcohol consumption [adjusted odds ratio (AOR) = 1.30; 95% CI: 1.04-1.62], households experiencing three (AOR = 1.78; 95% CI: 1.29-2.45) or four (AOR = 2.58; 95% CI: 1.86-3.58) climatic, biological, economic or conflict shocks in the prior 4 months compared to those experiencing less than 2 shocks, households with moderate (AOR = 1.25; 95% CI: 1.04-1.50) or high (AOR = 1.50; 95% CI: 1.22-1.85) water insecurity in the prior 4 weeks compared to those with no-to-marginal water insecurity, and child wasting (AOR = 1.22; 95% CI: 1.05–1.41).

These findings suggest that multisectoral interventions that reduce alcohol consumption among women, improve access to safe water services, manage malnutrition, and mitigate household shocks could reduce the burden of diarrhea among child under five in this region.

## Introduction

Diarrhea, the passage of three or more loose or liquid stools per day, is the third leading cause of death among children under five years of age globally [1]. Approximately 90% of these deaths occur in sub-Saharan Africa and South Asia [2]. Diarrhea also increases the risk of dehydration, appetite loss, malnutrition, electrolyte deficiencies, infectious disease, delayed physical growth, and cognitive impairments [1,3–5]. Malnutrition and diarrhea are interrelated, with each condition exacerbating the other, creating a cycle that severely compromises the long-term health and development of impacted children [6,7].

Previous studies in sub-Saharan Africa have identified factors associated with a higher risk of child diarrhea. These include younger child age [8–12], low maternal education [12–14], young maternal age [12,15], maternal unemployment [12,15], poor hand hygiene [16–18], open defecation [19–21], improper disposal of children’s stool [22,23], and households with more members [12,24,25]. Recent research also underscores the influence of behavioral, sociocultural, and socioeconomic factors on the prevalence of diarrhea in rural areas [26]. Given this, interventions that increase access to child-friendly sanitation infrastructure, change hand hygiene behavior and expand access to improved water services have been recommended to reduce the burden of child diarrhea [26,27]. A longitudinal study conducted in urban settings of developing countries confirmed the importance of sanitation as a major determinant of child diarrhea [28].

In sub-Saharan Africa’s arid and semi-arid lands (drylands), climate change has intensified droughts, significantly impacting livelihoods, household water, and food security [29–33]. Communities living in drylands experience food and water insecurity and predominantly rely on nomadic or semi-nomadic pastoralist livelihoods, increasing the risk of malnutrition [34].

Recurrent household climatic shocks in the region reduce agricultural productivity, damage critical infrastructure, and negatively impact health and well-being [15,35–37]. Additionally, limited access to improved water, sanitation, and hygiene services in the area increases exposure to vectors of diarrheal disease [3,32,38,39]. Moreover., households facing health shocks find themselves impoverished due to loss of income to facilitate access to quality health care and payment of health insurance [40,41].

While the distinctive conditions of drylands pose heightened risks for diarrhea, limited research investigates how environmental, socioeconomic, and behavioral factors intersect to influence diarrhea risk in these regions, particularly in Northern Kenya. To address this knowledge gap, we aim to assess the prevalence and risk factors of diarrhea among children under five in the drylands of Northern Kenya.

## Materials and methods

### Study area

This study was carried out as part of the USAID Nawiri program [42], to inform program design and adaptation. The study used data collected in Turkana County, which is in north-western Kenya and has seven sub-counties, 30 wards, and 156 sub-locations [43]. This region is characterized by low annual rainfall, high year-round temperatures, and a topographically varied landscape that ranges from semi-arid to arid. Traditionally, the Turkana people are nomadic pastoralists, relying on livestock and migrating in search of pasture to support their livelihoods. In addition to pastoralism, some communities practice agro-pastoralism, engaging in crop farming through irrigation along River Turkwel, and fishing on Lake Turkana [44].

### Study design

Caregivers with children under 36 months were eligible to participate. A multistage sampling strategy was used to ensure that findings were representative at the regional and sub-regional levels. In the first stage, the population was stratified based on administrative zones. Within each stratum, villages were randomly selected. Study staff identified all households in selected villages to create a sampling frame for eligible households. Households were then randomly selected from this frame and invited to participate. Further details on the study design are available in the study protocol [45].

### Data collection

Data were collected from May 2021 to September 2023 across six survey waves conducted every four months. Trained enumerators administered household and caregiver questionnaires to participating households. Study staff checked the integrity and validity of the data daily to ensure data quality.

### Dependent variable

Child diarrhea in the 2 weeks before the survey (yes/no) was based on caregivers’ self-report.

### Independent variables

Independent variables included distal, intermediate, and proximal factors, with wasting considered an additional independent variable given its bidirectional relationship with diarrhea (Figure 1). Distal factors included administrative and livelihood zones. Intermediate factors included head of household age (years), household head sex (man/woman), household wealth, number of household members, child age (months), child sex (male/female), child nutrition indicators, caregiver age (years), caregiver education (formal/no formal education), caregiver marital status (in union: currently living together; not in union: separated, divorced, widowed or single), and whether the household experienced shocks in the four months before the survey. Proximal factors included water, sanitation, and hygiene (WASH) conditions and behaviors. Caregivers’ handwashing after visiting the toilet in the last 24 hours (yes/no) and household practice of open defection (yes/no) were based on self-report.

**Figure 1.**
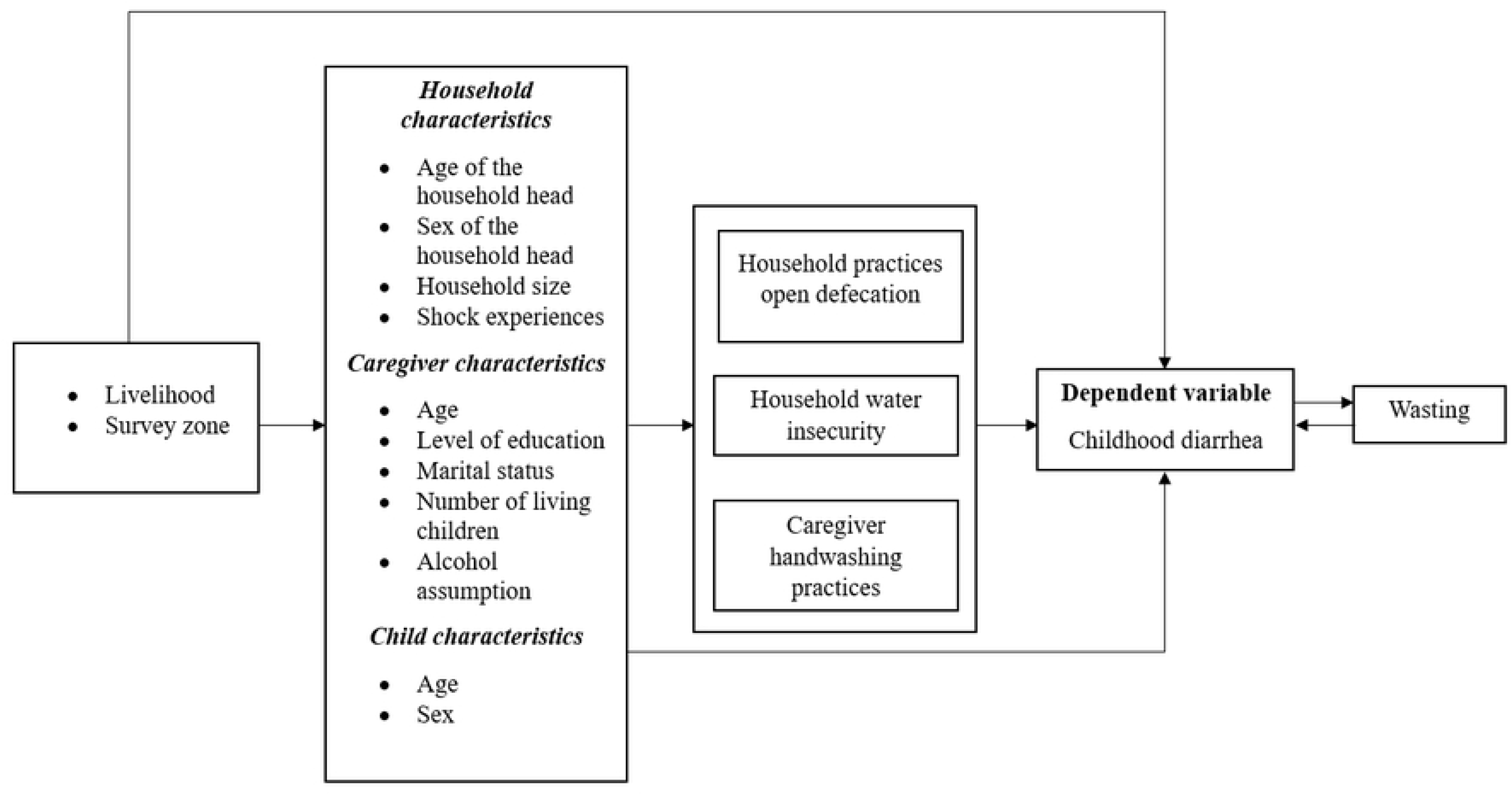
A conceptual framework for diarrhea risk factors among children under five.

The county was divided into four administrative zones (Central =1, North =2, South = 3 and West = 4) which comprised sub-counties: Central (Turkana Central and Loima), North (Turkana North and Kibish), South (Turkana East and Turkana South) and West (Turkana West). The livelihood zones in Turkana County were broadly categorized into four zones namely pastoral, agro-pastoral, fisher folks and urban/peri-urban. However, due to small sample sizes, the agro-pastoral, fisherfolk and urban/peri-urban zones were categorized as non-pastoral livelihood zone.

The household wealth was determined based on a factor analysis of household assets, housing materials, and access to utilities, and divided into tertiles (low, medium, and high). Household shocks were categorized into four types: climatic (excessive rains/flooding, variable rain/drought), biological (livestock/crop/human disease outbreak, crop pest invasion, weed outbreak, and severe illness), economic (loss of livelihood, increased prices in food/agricultural/livestock inputs, loss of land/rental property, youth unemployment, death of a household member, delay in food assistance, delay in other safety net programs, and fire) and conflict (theft/destruction of assets, theft of livestock, domestic violence, and community conflicts) and were binary variables. Further, the shock responses were summed up, and households were categorized as having experienced less than two, two, three, and four shocks.

Household water insecurity was measured using the 12-item Household Water Insecurity Experiences (HWISE) Scale [46]. Caregivers were asked to report how frequently in the prior 4 weeks they or others in their household experienced diverse water problems (0 = “never”, 1 = “rarely”, 2 = “sometimes”, 3 = “often” or “always”). Responses were summed and households were categorized as experiencing no-to-marginal (scores of 0-2), low (3-11), moderate (12-23), or high (24-36) water insecurity. Child wasting was assessed using weight-for-height/length Z-scores (WHZ < −2 SD), based on the 2006 World Health Organization (WHO) child growth standards [47].

## Data analysis

Descriptive analyses were performed to examine the weighted distribution of distal factors, household, caregiver, and child characteristics at Wave 1. We performed univariate analysis to assess the associations between the dependent variable (child diarrhea) and the independent variables. The independent variables with a *p-value* < 0.20 were retained and used to build the final multivariable logistic regression models. To identify salient risk factors of diarrhea, iterative modeling was used to account for the complex interrelationships between the different independent variables of interest, adjusting for either distal, intermediate, or proximal factors based on the conceptual framework in Fig. 1 [48]. Multivariable logistic regressions of child diarrhea and independent variables using generalized estimating equations with working correlation matrices were used to account for the within-subject correlation of repeated measurements across survey waves [49,50]. Only those variables with *p-value* < 0.05 were considered factors associated with diarrhea among children under five years. Adjusted odds ratios (AOR) and 95% confidence intervals (CIs) were obtained from the adjusted multivariable logistic regression models and interpreted for significant covariates. All statistical analyses were performed using Stata 18 (Stata Corp, College Station, TX USA). The survey proportion command in Stata was used to compare each period and potential factors. We performed a non-parametric Cochran-Armitage test for the trend in the prevalence across time using nptrend command and p-value. The xtgee command was used to perform univariate and multivariable logistic regressions.

## Ethical Considerations

The study was approved by the African Medical and Research Foundation Ethical and Scientific Review Committee (Amref ESRC P905/2020) and the National Commission for Science, Technology, and Innovation of Kenya. The institutional review boards at the African Population and Health Research Center and Research Triangle Institute International signed a reliance agreement. Informed consent was obtained from the legal guardians of participating children.

## Results

A total of 1,211 households with eligible children were included in this study and their baseline descriptive statistics are summarised in Table 1. Overall, 63.9%, lived in the pastoral livelihood zone. Around two-thirds (61.9%) of the households were headed by men, while females headed a third (38.1%). 44.6% and 30.6% of households experienced three and four shocks in the 4 months before the survey respectively. Among caregivers, the average age was 33 years, 86.4% had no formal education, 49.6% lived with 3 to 5 children, and 85.2% were in a marital union. Almost half of the children were boys (54.8%) with 21.8% of children wasted and 32.1% reported to have experienced diarrhea in the two weeks before the survey.

**Table 1.** Distribution of distal factors, household, caregiver, and child characteristics at baseline, Turkana County (N = 1,211)

| Characteristic | Weighted % |
| --- | --- |
| **Distal factors** |  |
| Livelihood zone |  |
| Pastoral | 63.9 |
| Non-pastoral | 36.1 |
| Administrative zone |  |
| Central | 21.0 |
| North | 25.6 |
| South | 24.1 |
| West | 29.2 |
| **Household characteristics** |  |
| Age of the household head (years) <sup>a</sup> | 39.5±13.4 |
| Sex of household head |  |
| Man | 61.9 |
| Woman | 38.1 |
| Household size <sup>a</sup> | 6.1±2.1 |
| Wealth tertile |  |
| Lowest | 45.6 |
| Middle | 33.8 |
| Highest | 20.6 |
| Shocks experienced in the last 4 months before the survey |  |
| Less than two | 4.2 |
| Two | 20.6 |
| Three | 44.6 |
| Four | 30.6 |
| **Caregiver characteristics** |  |
| Age (years) <sup>a</sup> | 33.1±10.2 |
| Education |  |
| Formal education | 13.6 |
| No formal education | 86.4 |
| Marital status |  |
| In union | 85.2 |
| Not in union | 14.8 |
| Number of living children |  |
| Less than 3 | 25.9 |
| 3-5 | 49.6 |
| 6 or More | 24.4 |
| Current alcohol consumer | 8.3 |
| ***Child characteristics*** |  |
| Sex |  |
| Male | 54.8 |
| Female | 45.2 |
| Age (months)<sup>a</sup> | 17.3±10.6 |
| Wasting (Weight-for-height/length z score < -2SD) |  |
| Yes | 21.8 |
| Experience of diarrhea in the 2 weeks before the survey |  |
| Yes | 32.1 |

<sup>a</sup> Mean ± standard deviation

The overall weighted prevalence of diarrhea and selected variables in each period were examined (**Table 2**). The data revealed that 32.1% of the children were reported to have experienced diarrhea in Wave 1. There was a noticeable decrease from 32.1% to 27.0% in one year, followed by a further steady decrease to Wave 4. Between Wave 4 to 5, there was a notable increase in the prevalence by approximately 2%, followed by a decrease of 9.1% to Wave 6. A similar trend was observed in the livelihood, administrative zones and child sex. A statistically significant trend in the prevalence of diarrhea was observed in the overall estimates and the livelihood zones. The prevalence of diarrhea in administrative zones and child age did not have a statistically significant trend.

**Table 2.**
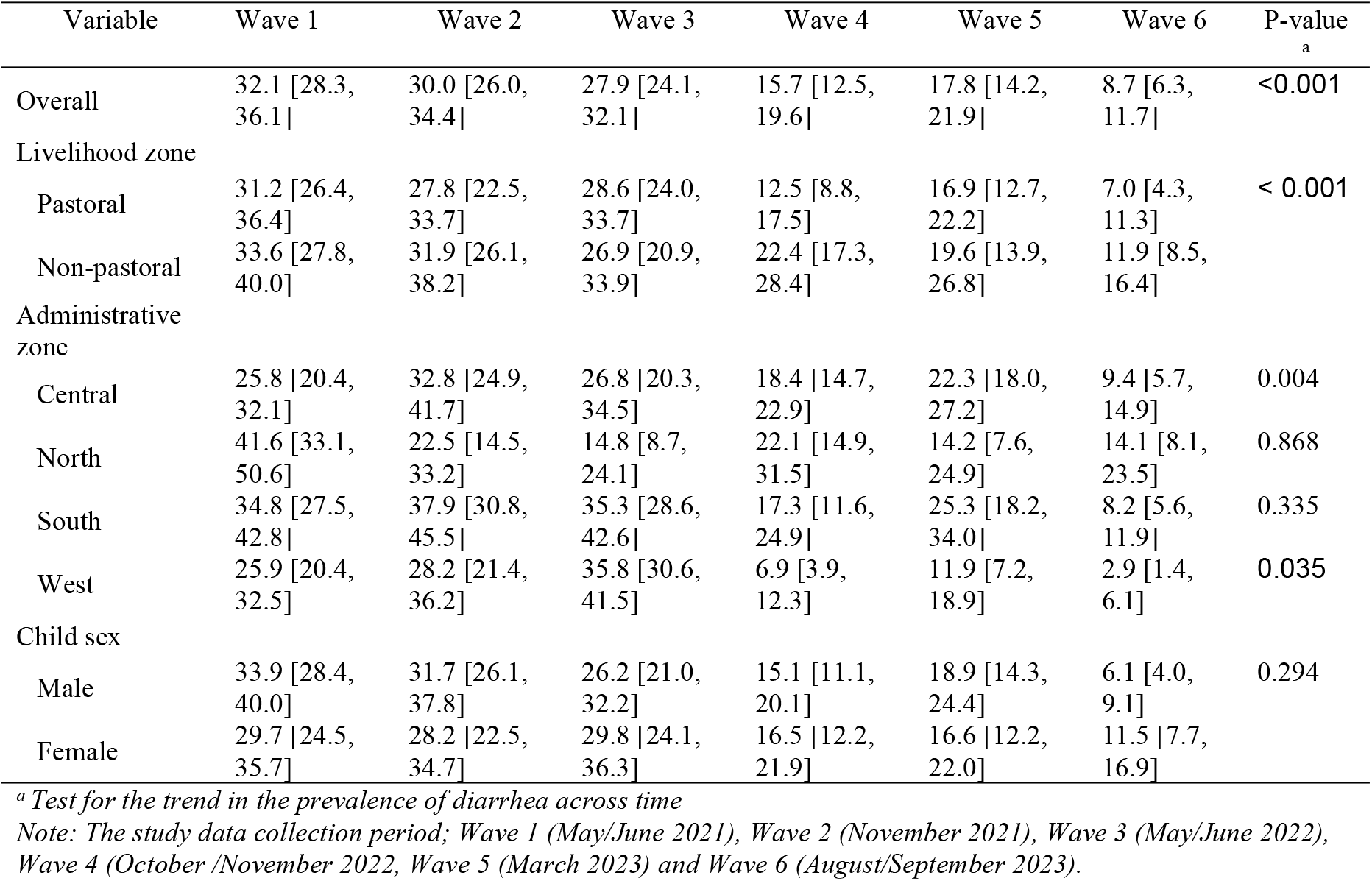
Weighted prevalence of diarrhea by livelihood zone, administrative zone, and child sex across survey waves (%, [95% CI]).

In bivariate analysis, the following factors were associated with diarrhea at *p*<0.2: distal factors (livelihood and administrative zones), intermediate factors (child age and sex, caregiver age, caregiver number of living children, caregiver alcohol consumption, head of household age, household shocks), proximal factors (household water insecurity, handwashing of caregiver after visiting toilet, household open defecation) and childhood wasting (**S1 Table**). After adjusting for potential confounders in multivariable models (**Table 3**), children with caregivers who consumed alcohol were found to have higher odds of diarrhea (AOR = 1.30; 95% CI: 1.04 - 1.62) compared to those whose caregivers did not. In addition, children in households that experienced three (AOR = 1.78; 95% CI: 1.29 - 2.45) or four (AOR = 2.58; 95% CI: 1.86 - 3.58) shocks had higher odds of diarrhea compared to those in households that experienced fewer than two shocks. Relative to children living in households with no-to-marginal water insecurity, those in households with moderate (AOR = 1.25; 95% CI: 1.04 - 1.50) or high-water insecurity (AOR = 1.50; 95% CI: 1.22 - 1.85) had higher odds of diarrhea. The analysis revealed that the households within the pastoral livelihood zone, children’s age, number of children living with the caregiver and handwashing practice by the caregiver were protective factors against diarrhea.

**Table 3.**
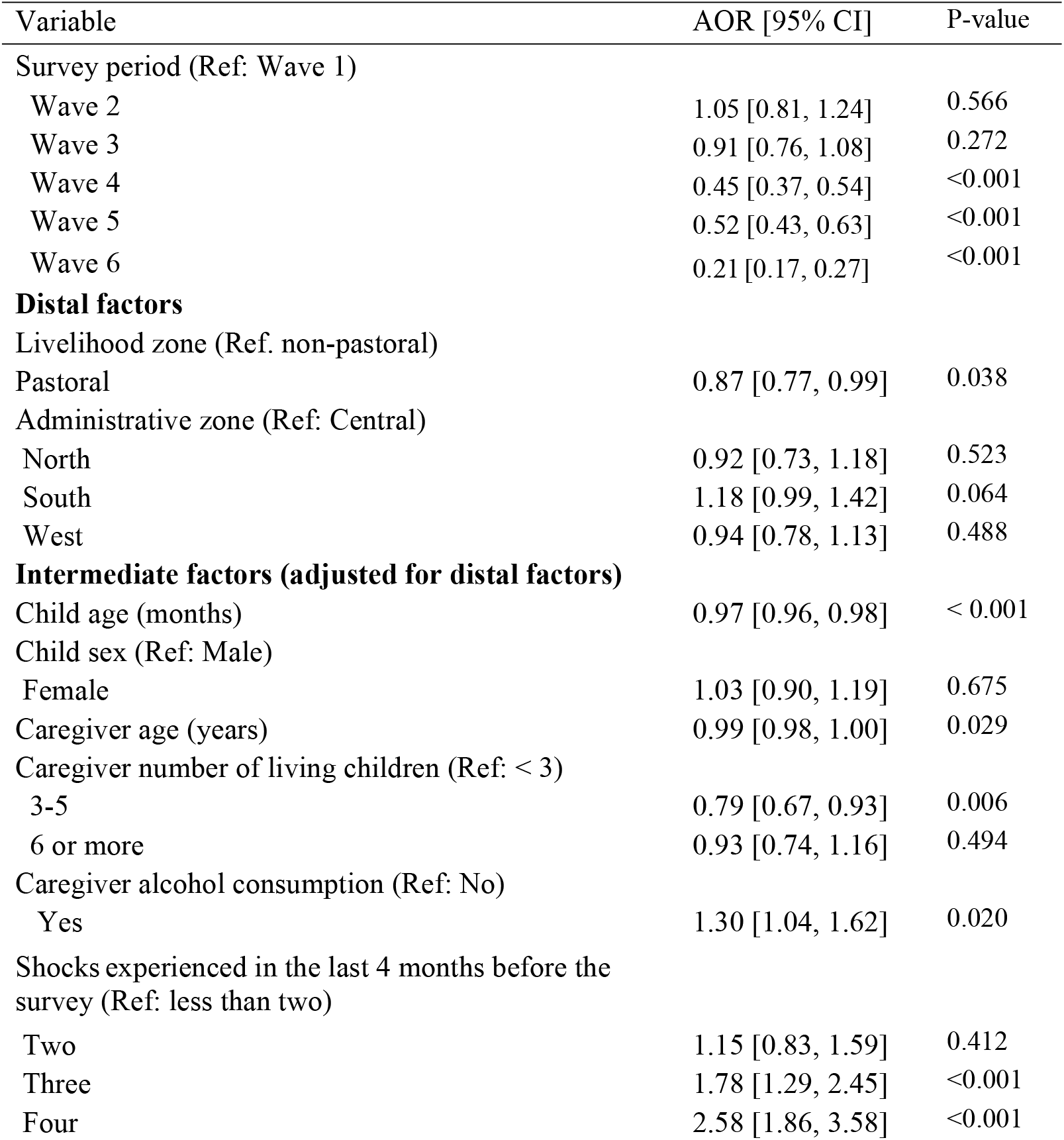
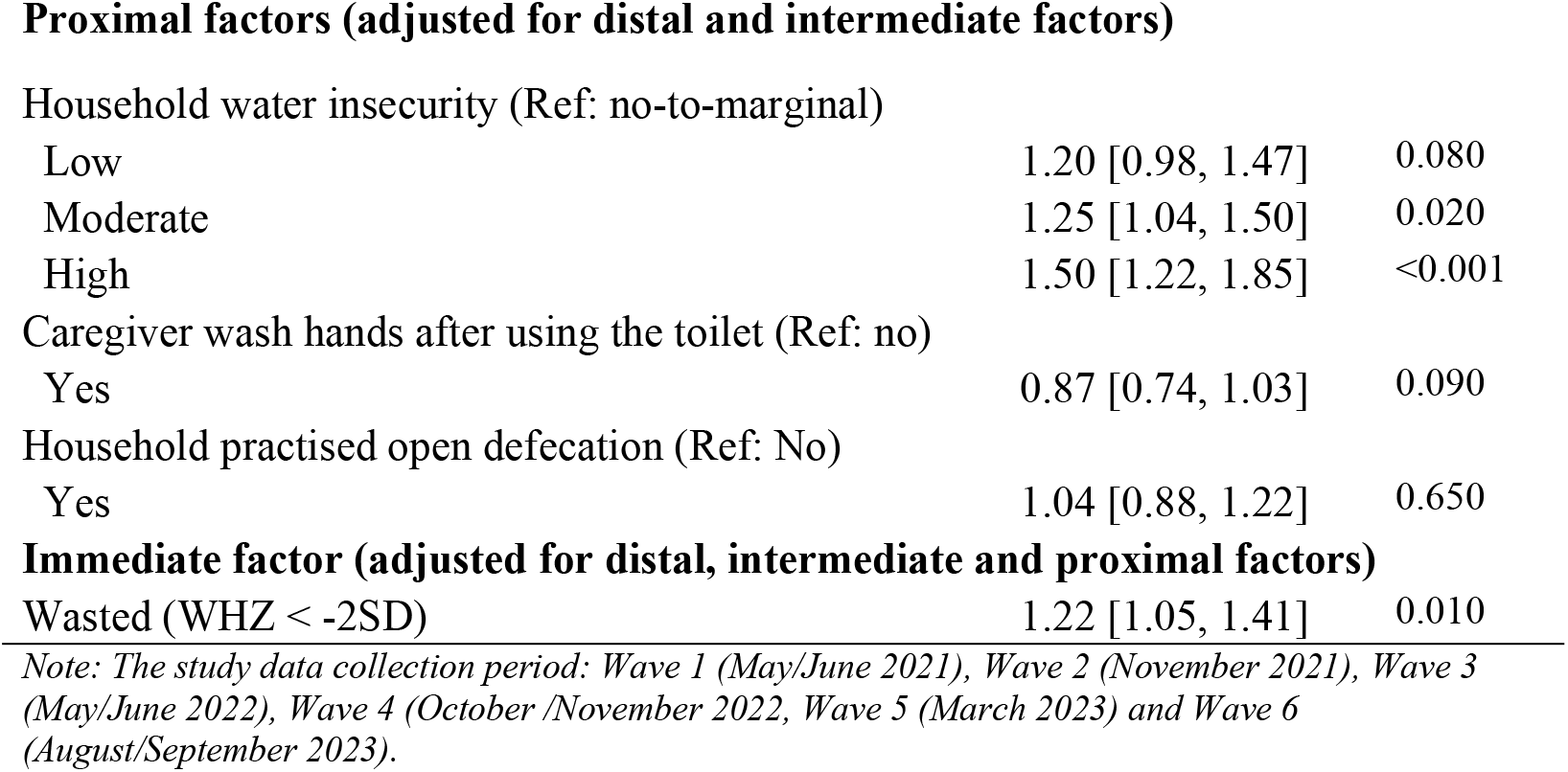
Factors associated with diarrhea among children under five in Turkana County, Kenya.

| Variable | AOR [95% CI] | P-value |
| --- | --- | --- |
| Survey period (Ref: Wave 1) |  |  |
| Wave 2 | 1.05 [0.81, 1.24] | 0.566 |
| Wave 3 | 0.91 [0.76, 1.08] | 0.272 |
| Wave 4 | 0.45 [0.37, 0.54] | <0.001 |
| Wave 5 | 0.52 [0.43, 0.63] | <0.001 |
| Wave 6 | 0.21 [0.17, 0.27] | <0.001 |
| **Distal factors** |  |  |
| Livelihood zone (Ref. non-pastoral) |  |  |
| Pastoral | 0.87 [0.77, 0.99] | 0.038 |
| Administrative zone (Ref: Central) |  |  |
| North | 0.92 [0.73, 1.18] | 0.523 |
| South | 1.18 [0.99, 1.42] | 0.064 |
| West | 0.94 [0.78, 1.13] | 0.488 |
| **Intermediate factors (adjusted for distal factors)** |  |  |
| Child age (months) | 0.97 [0.96, 0.98] | < 0.001 |
| Child sex (Ref: Male) |  |  |
| Female | 1.03 [0.90, 1.19] | 0.675 |
| Caregiver age (years) | 0.99 [0.98, 1.00] | 0.029 |
| Caregiver number of living children (Ref: < 3) |  |  |
| 3-5 | 0.79 [0.67, 0.93] | 0.006 |
| 6 or more | 0.93 [0.74, 1.16] | 0.494 |
| Caregiver alcohol consumption (Ref: No) |  |  |
| Yes | 1.30 [1.04, 1.62] | 0.020 |
| Shocks experienced in the last 4 months before the survey (Ref: less than two) |  |  |
| Two | 1.15 [0.83, 1.59] | 0.412 |
| Three | 1.78 [1.29, 2.45] | <0.001 |
| Four | 2.58 [1.86, 3.58] | <0.001 |
| Proximal factors (adjusted for distal and intermediate factors) |  |  |
| Household water insecurity (Ref: no-to-marginal) |  |  |
| Low | 1.20 [0.98, 1.47] | 0.080 |
| Moderate | 1.25 [1.04, 1.50] | 0.020 |
| High | 1.50 [1.22, 1.85] | <0.001 |
| Caregiver wash hands after using the toilet (Ref: no) |  |  |
| Yes | 0.87 [0.74, 1.03] | 0.090 |
| Household practised open defecation (Ref: No) |  |  |
| Yes | 1.04 [0.88, 1.22] | 0.650 |
| **Immediate factor (adjusted for distal, intermediate and proximal factors)** |  |  |
| Wasted (WHZ < -2SD) | 1.22 [1.05, 1.41] | 0.010 |

*Note: The study data collection period: Wave 1 (May/June 2021), Wave 2 (November 2021), Wave 3 (May/June 2022), Wave 4 (October /November 2022, Wave 5 (March 2023) and Wave 6 (August/September 2023).*

## Discussion

In this study, we found that caregiver alcohol consumption, household water insecurity, household shocks, and child wasting analysis were associated with higher odds of diarrhea in children. These findings align with prior studies and can help to inform policies and programs that aim to reduce the high burden of child diarrhea in Turkana County, Kenya.

Households in the drylands of sub-Saharan Africa suffer from frequent shocks, extended periods of poverty, and vulnerability, which hinder access to food, water and primary healthcare. Studies have shown that shocks often exacerbate existing vulnerabilities, particularly among marginalized communities that heavily depend on livestock [33,51]. Further, research indicates that these vulnerabilities adversely affect child nutrition, a key risk factor for childhood morbidity and mortality, as families struggle to maintain adequate food intake and access to clean water during crises [52–54]. Our study provides further evidence on the effects of household experience of shocks on the risk of childhood diarrhea in an arid land in Kenya.

Pastoral livelihood in sub-Saharan Africa increases vulnerability to shocks due to recurrent droughts and famines rendering many destitute and chronically dependent on food aid. Some households adapt to this situation by diversifying into activities that generate cash income, such as petty trade, crafting, weaving, and basketry. Research has shown that when pastoral households experience loss of livestock, children are particularly vulnerable to illnesses such as diarrhea and respiratory infections, primarily due to poor nutrition and limited access to primary healthcare [55,56]. Further, a study conducted in three pastoralist communities in northern Kenya provides evidence for the importance of a contextualized approach detailing how social environments that include endemic conflict produce variations in health [57]. The present study has shown that pastoral livelihood is a protective factor for childhood diarrhea contrary to the previous findings, opening a research question for further investigations in the future.

Our study revealed that children of caregivers who reported alcohol consumption had significantly higher odds of experiencing diarrhea. While caregiver alcoholism has been associated with persistent malnutrition in children under five [58], there is limited research directly connecting it to diarrhea. Qualitative data from Wave 1 of this longitudinal study suggested that alcohol use compromised caregivers’ abilities to provide adequate and hygienic care, likely contributing to the observed association [59].

Greater experiences of household water insecurity (which includes issues with water availability, accessibility, and sufficiency for diverse household uses) have been similarly positively associated with diarrhea risk among children under 5 in Cameroon [60], Ethiopia [61], and Nigeria [62]. Most studies, however, have only examined child diarrhea related to directly observable, supply-side water indicators, such as water source and quality [63,64]. Our findings demonstrate that experiential measures of water insecurity provide complementary information to traditional water metrics and make clear that water insecurity is an important modifiable risk factor that should be targeted to improve child health and well-being.

Malnutrition, particularly wasting, was also significantly associated with the odds of child diarrhea. Malnutrition undermines child development by depleting nutritional reserves and compromising the immune system, thereby increasing susceptibility to diarrheal infections [65]. Diarrhea, in turn, can exacerbate malnutrition, leading to a bidirectional relationship between these two conditions that ultimately increases the risk of child mortality [66–68]. Previous studies have associated childhood wasting with severe diarrhea episodes [69,70]; our findings further support wasting as a key risk factor for diarrheal infection.

Our study had several limitations. First, caregiver-reported child diarrhea is subject to recall bias. Further, we did not ask about the type of diarrhea observed (e.g., acute watery, acute bloody, or persistent), as each might have different health impacts. Systematic reviews have, however, noted a lack of evidence supporting the validity of instruments used in pediatric diarrheal disease trials [71]. Future research is needed to develop and validate better child diarrhea measures so that analyses can be disaggregated by diarrhea sub-type. Second, this longitudinal study was not originally designed to assess diarrhea as a primary outcome, although the large sample size reduces the risk of underpowered analyses.

## Conclusion

In this study of diarrhea among children under five in the drylands of Northern Kenya, we found that the prevalence of diarrhea decreased as children got older. Baseline surveys were conducted during the dry season, which may have also contributed to the prevalence of diarrhea observed at this time point. The findings on diarrhea prevalence disaggregated across survey waves can serve as useful referents for future targeted interventions in this region, particularly among non-pastoralists and households in the North zone, where the prevalence was highest. Key risk factors for diarrhea in Turkana County included caregiver alcohol use, household experience of shocks, household water insecurity, and child wasting.

A multisectoral approach is recommended to reduce the burden of diarrheal disease among children under five in this region. Such an approach should prioritize interventions that improve access to improved WASH services, address malnutrition prevention and management, and reduce the burden of household economic and climatic shocks. Lastly, there’s a need for intervention to reduce alcohol consumption among women as this may contribute towards reducing the burden of diarrhea among children under five years.

## Data Availability

The data underlying this study will be made available to researchers through the APHRC’s Microdata Portal (https://aphrc.org/microdata-portal/).

## Author contributions

FT, DA, VLF, CKL, AW, and ES designed the study. CW, DA, BM, EA, HO, SE, and GC implemented fieldwork. CW, ES, FT, and EK-M provided study oversight. BM, JDM, and CW analyzed the data. BM prepared the first draft of the manuscript and incorporated comments from co-authors. All authors contributed to the revision of the manuscript and approved the final version.

## Supplementary Tables

**S1 Table**. Bivariate logistic regression analysis of factors associated with diarrhea among children under five years of age

